# Mutations in *CSRP3/MLP*, a Z-disc associated gene are functionally associated with dilated cardiomyopathy in Indian population

**DOI:** 10.1101/2021.11.05.21265852

**Authors:** Prerna Giri, Ritu Dixit, Ashok Kumar, Bhagyalaxmi Mohapatra

**Affiliations:** Cytogenetics Laboratory, Department of Zoology, Institute of Science, Banaras Hindu University, Varanasi-5, Uttar Pradesh, India; Department of Pediatrics, Institute of Medical Sciences, Banaras Hindu University, Varanasi, Uttar Pradesh, India

**Keywords:** DCM, Missense, Cardiomyopathy, *CSRP3*, LIM-domain protein, Zinc-finger

## Abstract

CSRP3 is a LIM domain containing protein, known to play an important role in cardiomyocyte development, differentiation and pathology. Mutations in *CSRP3* gene are reported in both dilated and hypertrophic cardiomyopathy (DCM and HCM), however, the genotype-phenotype correlation still remains elusive. To investigate the pathogenic potential of *CSRP3* variants in our DCM cohort, we have screened 100 DCM cases and 100 controls and identified 3 non-synonymous variations, of which two are missense variants viz., c.233 GGC>GTC, p.G78V; c.420 TGG>TGC, p.W140C, and the third one is a single nucleotide polymorphism (SNP) c.46 ACC>TCC, p.T16S. These variants were absent from 100 control individuals (200 chromosomes). *In vitro* functional analysis has revealed reduction of CSRP3 protein level in stably-transfected C2C12 cells with p.G78V or p.W140C variants. Immunostaining demonstrates both cytoplasmic and nuclear localization of the wild-type protein, however variants p.G78V and p.W140C cause obvious reduction in the cytoplasmic expression of CSRP3 protein which is more pronounced in case of p.W140C. Disarrayed actin cytoskeleton was also observed in mutants. Besides, the expression of target genes namely *Ldb3, Myoz2, Tcap, Tnni3* and *Ttn* are also downregulated in response to these variants. GST-pulldown assay has also showed a diminished binding of CSRP3 protein with α-Actinin due to both variants p.G78V and p.W140C. Both 2D, 3D-modeling have shown confirmational changes. Most *in silico* tools predict these variants as deleterious. Taken together, all these results suggest the impaired gene function due to these deleterious variants in CSRP3, implicating its possible disease causing role in DCM.

## 1. Introduction

Dilated cardiomyopathy (DCM) is one of the most prevalent types of idiopathic cardiomyopathy, characterized by left ventricular wall thinning and enlargement of chamber along with severe systolic dysfunction (Richardson et al., 1996). Although the etiology of DCM is not well known, approximately 30-40 % of DCM cases have a genetic basis (Grunig et al. 1998, Kelly et al. 1994, Mestroni et al. 1995 and Keeling et al. 1995). More than 60 genes have been identified in different subcellular compartments, such as sarcomere, cytoskeleton, sarcolemma, nucleus, nuclear lamina, mitochondria, plasmalemma and Z-disc to be associated with the disease (Giri et al.,2021). Among these, a gene encoding cysteine and glycine-rich protein3 (CSRP3), which is also known as cardiac LIM domain protein (CLP) or muscle LIM protein (MLP) (HGNC:24722; Chr. Location: 11p15.1), has been associated with variety of cellular process i.e., growth, differentiation, contractility, maintenance of cyto-architecture and signal transduction (Kadrmas and Beckerle, 2004; Vafiadaki et al., 2015). The protein is primarily known for its role in striated muscle development, differentiation and pathology (Arber et al. 1994,Kong et al 1997), which expresses specifically in slow skeletal muscle and cardiac tissues. The first evidence of implication of *CSRP3* in cardiac disease was reported by Zolk et al., 2000, which showed a significant reduction in CSRP3 protein levels in human failing hearts of both dilated and ischemic cardiomyopathy patients. The CSRP3 deficient mice displayed severely impaired cardiac function exhibiting characteristic features of DCM followed by progressive heart failure (Arber et al., 1997). On the other hand, knock-in mouse models using either W4R or C58G point mutations caused hypertrophic cardiomyopathy and heart failure (Knöll et al., 2010; Ehsan et al., 2018). Ablation of *CSRP3* gene in mouse model showed overexpression of a gap junctional protein N-RAP and impaired myofibrillar structure and function leading to DCM phenotype (Ehler et al., 2001). Further, another MLP-KO study in mouse depicted decreased mitochondrial content in the myocardium and consequent energy depletion leading to decreased cardiac function (van den Bosch 2005). However, mouse model over-expressing MLP has been shown to display normal cardiac characteristics (Kuhn et al. 2012).

The CSRP3/MLP encode a 194 amino acid long protein (Fung et al. 1996 and Knoll et al. 2002), which consists of two LIM domains viz., LIM1 and LIM2 and next to each LIM domain is neighbouring glycine rich (GR) modules (Weiskirchen et al. 1995). Both the LIM domains are highly conserved domains possessing two Zinc (Zn) finger motifs each (Zheng and Zhao 2007). All the three cysteine rich proteins (CSRP1, CSRP2 and CSRP3) are highly conserved indicating their evolutionary significance. CSRP3 has been primarily localized to sarcomeric Z-disc, considered as a mechanical stretch-sensor, interacting with telethonin (TCAP), (Knoll et al., 2002),α-actinin (Mohapatra et al 2003; Gehmlich et al., 2004), CARP (Lange et al., 2016) and ZASP/Cypher (Kimura 2015). The protein is also localized to costamere, interacting with zyxin and spectrin (Sadler et al., 1992; Flick and Konieczny 2000), while in intercalated disc it interacts with N-RAP (Ehler et al., 2001). Similarly in nucleus, it interacts with several transcription factors viz., GATA4, SRF, MyoD and MRF4 (Buyandelger et al., 2011). CSRP3 is present in two forms, oligomeric and monomeric (Boateng et al., 2007). Oligomeric forms are found in cytoskeleton and monomeric forms in nucleus. The CSRP3 also plays an important role in sarcomere assembly and cytoskeleton integrity which is accomplished by its binding to α-actinin, a protein implicated in regulation of actin organization which is also involved in cross-linking actin filaments (Dubreuil, 1991).

CSRP3 has been implicated in muscle differentiation, due to its interaction with basic helix-loop-helix transcription factor (bHLH). To date, more than 25 mutations in human *CSRP3* encoding MLP protein have been reported to be associated with both dilated (DCM) as well as hypertrophic cardiomyopathy (HCM) (Lipari et al., 2020). MLP W4R mutation was the first reported mutation in DCM patients (Knoll et al., 2002). However, the variant W4R has shown both DCM and HCM phenotype (Knoll et al.,2010). Linkage analysis of large multigenerational families implicated causation of HCM due to mutations in *CSRP3* (Geier et al., 2008). However, the causative association of *CSRP3* gene with either HCM or DCM remain unclear.

In the present study, we have screened *CSRP3* in 100 patients with idiopathic DCM in Indian population and report three misssense variations by Sanger’s sequencing. *In vitro* functional studies, supported by bioinformatic analysis revealed impaired gene function.

## 2. MATERIALS & METHODS

### 2.1 Clinical Evaluation and Enrolment of Patients

A cohort of 100 probands including 7 familial cases (45 males and 55 females) with idiopathic DCM (median age at diagnosis-11 years) and 100 ethnicity matched healthy control individuals (median age 3.7 years) from Department of Pediatric Medicine and Department of Cardiology, S S Hospital, Institute of Medical Sciences, Banaras Hindu University, were recruited for this study. All study subjects were evaluated by physical examination, chest radiography, MRI, ECG and 2D-echocardigraphy. The diagnosis of DCM was based on the LV size and function (i.e. LV fractional shortening <25%, an ejection fraction of <45%). The study was approved by the Institutional Ethics Committee.

### 2.2 Mutational Analysis of MLP/CSRP3

For genetic investigations, peripheral venous blood samples (3-5ml) were collected from the patients, their available relatives and healthy control individuals, after written informed consent. Genomic DNA was extracted according to standard ethanol precipitation protocol. Primers were designed in the exon-intron boundaries of human *CSRP3* gene (NM_003476.4) and all the coding exons were amplified by polymerase chain reaction (PCR). After purification using Exonuclease I and Shrimp Alkaline Phosphatase (USB Products, Affymetrix, Inc, OH, USA), the PCR products were Sanger sequenced using Big Dye Terminator sequencing kit (ABI, MA, USA) as per manufacturer’s protocol, with the help of ABI PRISM 3130XL genetic analyzer. Finch TV software (http://www.geospiza.com/ftvdlinfo.html, Geospiza) was used to analyze the chromatograms. Each identified variant was confirmed by re-sequencing of freshly amplified PCR products of same amplicon from the DNA of respective subjects. Novelty of the sequence variants was checked by searching dbSNP (https://www.ncbi.nlm.nih.gov/snp/), ClinVar (https://www.ncbi.nlm.nih.gov/clinvar/), 1000 Genome (https://www.internationalgenome.org/) and Genome aggregation database (gnomAD) (https://gnomad.broadinstitute.org/).

### 2.3 In silico characterization of non-synonymous variants

#### 2.3.1 Phylogenetic conservation of amino acid residues in MLP protein

The reference genomic DNA sequence, mRNA sequence and protein sequence of *CSRP3* were taken from Gen-Bank (https://www.ncbi.nlm.nih.gov/genbank/) and Protein database (https://www.ncbi.nlm.nih.gov/protein/) for all the *insilico* analyses. Multiple sequence alignment of CSRP3 protein (NP_003467.1) homologs across different species was performed using the ‘HomoloGene’ feature of NCBI to evaluate the conservation of substituted amino acids.

#### 2.3.2 Prediction of pathogenic potential of variants identified in CSRP3/MLP

The disease-causing potential of non-synonymous variants identified in *CSRP3* was predicted using VarCards (http://159.226.67.237/sun/varcards/) which incorporates more than 10 *in silico* predictive algorithms e.g., PolyPhen-2 (http://genetics.bwh.harvard.edu/pph2/), SIFT (http://sift.jcvi.org/www/SIFT_enst_submit.html), Mutation Taster (http://www.mutationtaster.org), I-Mutant (http://folding.biofold.org/i-mutant/i-mutant2.0.html), FATHMM (http://fathmm.biocompute.org.uk/fathmmxf/index.html), VEST3 (https://karchinlab.org/apps/appVest.html) and M-CAP (http://bejerano.stanford.edu/mcap/), PROVEAN (Protein Variant Effect Analyzer, http://provean.jcvi.org/index.php), CADD (https://cadd.gs.washington.edu/), and SNAP^2^ etc (Screening of Non Acceptable Polymorphism, https://rostlab.org/services/snap2web/).

#### 2.3.3 Effect of non-synonymous variants, identified in CSRP3/MLP, on secondary and tertiary structure of protein

The secondary structure of the wild type and mutant (T16S, G78V and W140C) CSRP3 protein was predicted using PsiPred (http://bioinf.cs.ucl.ac.uk/psipred/) prediction tool. To further, elucidate the effect of non-synonymous variants on the overall protein structure, three-dimensional homology modeling of MLP protein (both wild type and mutants) was performed by utilizing the N terminal (PDB ID: 2O10_A) and C terminal available structures (PDB ID: 2O13_A) using AIDA server (Ab Initio Domain Assembly). The structure of wild type and mutant MLP protein modelled was subjected to energy minimization using YASARA sever for further analysis. For visualizing the differences in the modeled structures across wild type and mutant MLP protein, the two structures were aligned through UCSF Chimera was used for. The Root Mean Square Deviation (RMSD) values were calculated for each of the aligned structures.

#### 2.3.4 Assessment of physico-chemical properties

The changes in the physico-chemical properties (hydrophobicity, % accessible residues, relative mutability, α-helix, total β-strand) of CSRP3 protein due to non-synonymous variants were computed using the ProtScale (https://web.expasy.org/protscale/) server available at ExPasy portal. The profile for each of the physico-chemical property was obtained and compared across wild type and mutant CSRP3 protein to predict the effect of identified variants.

### 2.4 In vitro characterization of the non-synonymous variants

#### 2.4.1 Cloning of wild type CSRP3/MLP and Site Directed Mutagenesis

The wild type (WT) *CSRP3* was cloned into pcDNA3.1/V5-His TOPO TA (Invitrogen, Carlsbad, CA, USA) vector (as per the manufacturer’s protocol). The fidelity of the *CSRP3/MLP* construct was confirmed through DNA sequencing. WT-*CSRP3* was used as template for preparing mutant constructs of G78V-*CSRP3* and W140C-*CSRP3* by site directed mutagenesis using Quick change II XL site directed mutagenesis kit (Agilent Technologies Inc., Santa Clara, CA, USA) through megaprimer method as per manufacturer’s instructions.

#### 2.4.3 Expression of MLP in Cultured Mouse Myoblasts (C2C12 cell line)

In order to perform immunostaining for visualizing cellular localization and cytoskeletal assembly of WT and mutant CSRP3 (G78V, W140C), stable C2C12 cells expressing these WT and mutant CSRP3 were established. These cells were seeded in each well on glass coverslips in six well culture plates and grown in Dulbecco’s modified Eagle’s medium (DMEM, Gibco, Life technologies corp., NY, USA), supplemented with 10% Fetal Bovine Serum (FBS, Gibco, Life technologies corp., NY, USA) were incubated in 37°C at 5% CO_2_, overnight. Next day, cells (at 40-45% confluency) were transfected with 500 ng of either wild-type or mutant *CSRP3*-plasmid DNA, using FuGENE 6 transfection reagent (Promega Corp., IN, USA). After 48 hours of transfection, C2C12 cells were washed with 1X PBS twice and immediately fixed with 4% paraformaldehyde (PFA) for 15 minutes, followed by permeabilization with 0.5% Triton X (diluted in 1X PBS). After blocking with 5% non-fat dry milk (in 1X PBS) for 2 hours, cells were incubated with 1:500 dilution of primary antibody (Anti-His antibody, Sigma-Aldrich, MO, USA), overnight at 4°C, washed with 1X PBS 5-6 times, followed by incubation with Alexa Fluor 488 goat anti-mouse IgG (Molecular Probes, OR, USA) for 2 hours at room temperature. The cells were then washed extensively with 1X PBS (5-6 times), cytoskeleton was stained with Phalloidin (Sigma-Aldrich, MO, USA) and nuclei were stained with DAPI, mounted with mounting media and imaging was carried out using Zeiss LSM 510 Meta, Laser Scanning Confocal microscope, further analysed with ZEN 12 and LSM softwares and assembled using Adobe Photoshop software.

#### 2.4.4 Estimation of protein expression through Western Blotting

With a view to estimate the expression level of MLP protein in wild type versus mutants, C2C12 cells were seeded in six well plate and then transfected with 500 ng each of wild type and mutant (G78V and W140C) *CSRP3* constructs at 50% confluency of cells. Forty-eight hours post transfection, cell-lysates were prepared in RIPA buffer (50 mM Tris-HCl, pH 8.0, 150 mM NaCl, 0.1% Triton X, 0.5% sodium deoxycholate, 0.1 % SDS) with 1mM sodium orthovanadate, 1mM NaF, and protease inhibitor tablets (Roche, Basel, Switzerland). After dilution with Laemmeli’s sample buffer, the lysates were separated on 10 % polyacrylamide SDS gel and proteins were transferred to PVDF membrane (Bio-Rad Laboratories Inc, CA, US). The PVDF membrane was blocked for 2 hours in 5 % dry skimmed milk at room temperature, followed by overnight incubation with anti-His antibody (Sigma-Aldrich, MO, US) at 4°C, the blot was washed with PBST (0.1 % Tween-20 diluted in 1X PBS) six times for 10 mins each. The blot was then incubated with HRP conjugated goat anti-mouse IgG antibody (Genei, Merck Specialties Pvt. Ltd., Darmstadt, Germany) followed by washing with PBST (6X, 5 min each) and detected with ECL detection kit (GE Healthcare, IL, US).

#### 2.4.5 Real-Time Quantitative PCR (qPCR) Assay

The C2C12 cells were grown in a six well plate and at 40-45% confluency, both wild-type (pcDNA3.1-*CSRP3*-WT) and mutant (pcDNA3.1-*CSRP3*-G78V & pcDNA3.1-*CSRP3*-W140C) constructs were transfected (1ug/well) in these cells. Total cellular RNA was isolated at 48 hours post transfection using TRIzol™ reagent (Ambion, Inc, TX, US). All the RNA samples were digested with DNAse I (Thermo Fischer Scientific, MA, US) to remove genomic DNA contamination. RNA quality was checked on 1% agarose gel and further quantified using a NanoDrop1000 spectrophotometer. The cDNA was synthesized from 2 ug of DNAse I-digested RNA using Random hexamers and ‘Revertaid First Strand cDNA synthesis kit’ (Fisher Scientific, MA, US) as per manufacturer’s protocol.

The real time qPCR assays were performed to estimate the expression of *Tcap, Ttn, Tnni3, Myoz2, Ldb3* across wild type and mutant *CSRP3* transcripts using Sybr green (Puregene, Genetix Biotech Asia Pvt. Ltd., DL, IN) in a 7500 Real Time PCR System (Applied Biosystems, MA, US) in triplicate to compare the effects of wild-type and mutants (G78V, W140C) *CSRP3*. The expression data was normalized using the house keeping gene, β-actin as an internal control. All the data were represented as fold change of expression of mutant as compared to wild type, with standard error of mean. The p value was considered statistically significant when less than 0.05.

#### 2.4.6 Protein-Protein Interaction: GST Pull Down Assay

The pGEX-4T1 vector was used for GST pull down assay. The pcDNA3.1/V5-His *CSRP3* wild-type clone was double digested with BamH I and Xho I restriction enzymes and the *CSRP3* insert was eluted from the gel. Simultaneously, the pGEX-4T1 vector was also double digested with BamH I and XhoI and the linearized vector was gel-eluted. The linearized pGEX-4T1 vector was ligated with *CSRP3* insert, overnight at 16°C. The ligated product was transformed in DH5-alpha cells and the wild-type plasmids were isolated and digested with BamH I and Xho I to confirm the presence of *CSRP3* insert. The clone was further confirmed by Sanger sequencing. Mutant constructs expressing GST-*CSRP3*-G78V and GST-*CSRP3*-W140C fusion protein were obtained through site-directed mutagenesis using Quick change II XL site-directed mutagenesis kit (Agilent Technologies, CA, US) as per manufacturer’s instructions.

Both wild type and mutant GST-CSRP3 fusion protein were transformed in BL21-DE3 cells and induced using 0.1mM IPTG (Sigma Aldrich, MO, US) at their log phase (O.D. 0.5 at 600 nm wavelength) for 8 hours. Following induction, the bacterial cells were pelleted down, washed with 1X PBS, further resuspended in bacterial lysis buffer (2 ml bacterial lysis buffer was used per 100 ml of bacterial culture for resuspension) and incubated in ice with continuous shaking for 1 hour. Triton X-100 was added to cell lysate at a final concentration of 0.5 % followed by centrifugation at 13,000 rpm for 30 mins and the supernatant containing the bait protein was transferred to a fresh tube. Glutathione Sepharose 4B (GE Healthcare, IL, US) was used for immobilization of fusion proteins and GST protein. To over-express prey protein ACTN2, 10 ug of pCMV-*ACTN2*-c-myc was transfected in 100 mm culture dish seeded with C2C12 cells. Forty-eight hours, post transfection, protein lysate was prepared and incubated with GST beads overnight for pre-clearing once, following which mixture was centrifuged and supernatant containing pre-cleared lysate was collected. Equal amount of this lysate containing ACTN2 was added to GST-*CSRP3*-WT, GST-*CSRP3*-G78V, GST-*CSRP3*-W140C and GST protein bound slurry and incubated for overnight at 4°C. Bait-prey-bead complex (GST-CSRP3-WT-ACTN2, GST-CSRP3-G78V-ACTN2, GST-CSRP3-W140C-ACTN2) was centrifuged and bead was collected, washed with GST washing buffer II and incubated on ice for 10 min with shaking followed by centrifugation. After washing (6x), 60 μl of GST binding buffer was added into the bead complex, incubated for 15 mins at RT and then the bait-prey complex was eluted. The bait-prey protein complex was separated on 10 % SDS Gel, followed by Western blotting. Blot was probed with anti-CSRP3 antibody (Abcam, Cambridge, UK) for detection of CSRP3/MLP protein (bait protein) and anti-c-Myc tag (DHSB, IA, US) antibody for detection of ACTN2 protein (prey protein).

## 3. Results

### 3.1 Identification of Genetic Variants in MLP

Sanger’s sequencing of coding region and exon-intron boundaries of CSRP3/*MLP* in 100 probands revealed a total of 3 non-synonymous variants (Figure 1.A and C), two of which were missense variants (c.233 GGC>GTC, p.G78V; c.420 TGG>TGC, p.W140C) and the third one (c.46 ACC>TCC, p.T16S) was a single nucleotide polymorphism (MAF 0.12). None of these variants were present in 100 ethnicity-matched control individuals (200 chromosomes). Two of these variants (p.T16S andp.W140C) were novel, while the variant p.G78V (rs963128995) was already reported (Table 1). All the three variants were submitted in ClinVar database, the RCV numbers of the submitted variants are as follows: c.46 ACC>TCC, p.T16S (RCV000710022.1); c.233 GGC>GTC, p.G78V (RCV000710024.1); c.420 TGG>TGC, p.W140C (RCV000710027.1). Minor allele frequency (MAF) for these variants and the clinical phenotype of patients are shown in Table 1.

**Table 1.** Table showing clinical characteristics, ClinVar submission ids, domains to which variants are localized and the pathogenic potential of non-synonymous variants identified in *CSRP3*.

| Nucleotide Change | Amino Acid Change | No. of Case (s) (MAF) | No. of Control (s) | dbSNP | Left Ventricular Ejection Fraction (%) | Left Ventricular Internal End Diastolic Diameter (mm) | Domain | Pathogenicity<br>(*In silico*) |  |  |  |  |  |  |  |  |  |
| --- | --- | --- | --- | --- | --- | --- | --- | --- | --- | --- | --- | --- | --- | --- | --- | --- | --- |
|  |  |  |  |  |  |  |  | Polyphen2HDIV<br>(Cut off=0.05) | Mutation Taster | SIFT<br>(Cut off=0.05) | VEST3<br>(Cut off=0.05) | FATHMM<br>(Cut off=0.05) | M_CAP | PROVEAN | CADD | I-Mutant | SNAP<sup>2</sup><br>(-100 to 100) |
| c.46<br>ACC>TCC | p.T16S | 24/100<br>(0.12) | 0/100 | Novel<br>(RCV000710022.1) | - | - | First LIM Domain | Benign<br>0.035 | Disease Causing<br>(1) | Tolerable (0.614) | Damaging<br>(0.607) | Damaging<br>(-2.79) | Tolerable<br>(0.021) | Tolerable<br>(-0.11) | Tolerable<br>(9.142) | Decreased Stability | Neutral<br>(-63) |
| c.233<br>GGC>GTC | p.G78V | 1/100<br>(0.005) | 0/100 | rs963128995<br>(RCV000710024.1) | 26 | 68 | Glyine Rich Module | Probably Damaging (1) | Disease Causing<br>(1) | Damaging(0.001) | Damaging<br>(0.894) | Damaging<br>(-3.14) | Damaging<br>(0.207) | Damaging<br>(-7.72) | Damaging<br>(33) | Decreased Stability | Effect<br>(62) |
| c.420<br>TGG>TGC | p.W140C | 1/100<br>(0.01) | 0/100 | Novel<br>(RCV000710027.1) | 16 | 72 | Second LIM Domain | Probably Damaging (1) | Disease Causing<br>(1) | Damaging(0.00) | Damaging<br>(0.986) | Damaging<br>(-2.81) | Damaging<br>(0.392) | Damaging<br>(-11.23) | Damaging<br>(33) | Decreased Stability | Effect<br>(76) |

Nomenclature refers to *CSRP3* reference sequence (NM\_003476.4). For cDNA positions, +1 refers to the base A of first codon ‘ATG’; for protein (NP\_003467.1) positions +1 refers to amino acid coded by first codon ‘ATG’ Abbreviations: MAF -minor allele frequency; dbSNP –single nucleotide polymorphism database; RCV-ClinVar database accession number of submitted varaints

**Figure 1.**
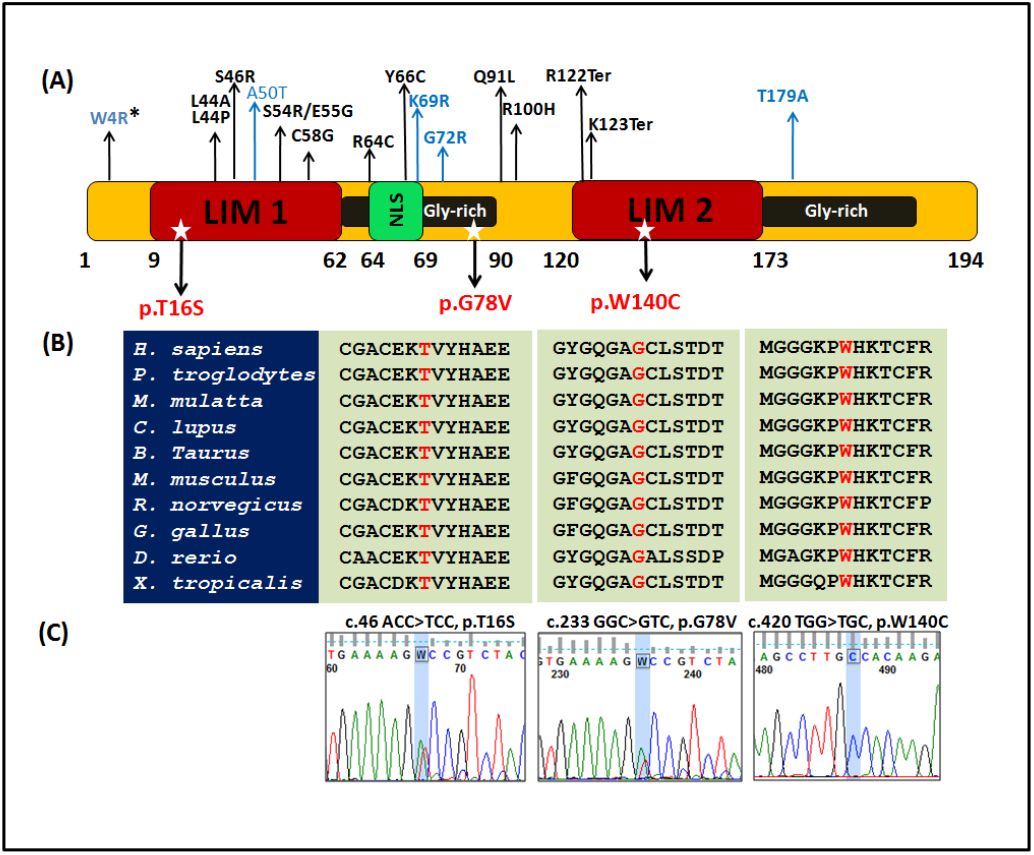
Schematic representation of *CSRP3* variants and their phylogenetic conservation. (A) Diagrammatic representation of CSRP3 protein (194 amino acid) showing different domains and modules of CSRP3 protein, viz., LIM1 (9-62 amino acids), LIM2 (120-173 amino acids) domain, gly-rich modules (63-90 amino acids) and the NLS (Nuclear Localization Signal) which is located within amino acid residues 64-69. Position of identified non-synonymous are marked with star while previously published DCM and HCM associated non-synonymous variants are marked with blue and black arrows respectively. (B) Phylogenetic conservation of CSRP3 protein across different species showing substituted amino acids (highlighted in red). (C) Sequence chromatogram of non-synonymous variants c.ACC>TCC, c.233 GGC>GTC and c.420 TGG>TGC, p.W140C respectively.*W4R non synonymous variant has been reported to be associated with both DCM and HCM

The patient harboring non synonymous variant p.G78V (MAF 0.005) had a highly reduced LVEF=26% and increased LVIDd=68 mm. The p.W140C (MAF 0.01) was a homozygous substitution missense variation and the patient harboring this variant had a LVEF=16% and LVIDd= 72 mm. The LVEF and LVIDd values indicate that left ventricular systolic function was severely impaired. Other relatives of this patient were also diagnosed with DCM, however died at relatively younger age and could not be tested for this genetic mutation. Apart from these two non-synonymous variants, variant p.T16S that has been identified in 24 cases (MAF 0.12), is a novel non-synonymous single nucleotide polymorphism (SNP) (Table. 1). Since the minor allele frequency (MAF 0.12) of this polymorphism was relatively higher than the other two non-synonymous variants, this variant was not selected for further in vitro functional characterization. The two rare variants were characterized for their effect on the overall function of the protein using appropriate *in silico* and *in vitro* techniques.

### 3.2 *Phylogenetic conservation of* CSRP3 *across species*

A cross species multiple sequence alignment of CSRP3 protein across several species viz., *Homo sapiens* ([NP_003467.1](https://www.ncbi.nlm.nih.gov/entrez/viewer.fcgi?db=protein&id=4502893)), *Pan troglodytes* ([XP_528813.2](https://www.ncbi.nlm.nih.gov/entrez/viewer.fcgi?db=protein&id=114636532)), *Macaca mulatta* ([XP_001095430.1](https://www.ncbi.nlm.nih.gov/entrez/viewer.fcgi?db=protein&id=109107042)), *Canis lupus familiaris* ([XP_852328.1](https://www.ncbi.nlm.nih.gov/entrez/viewer.fcgi?db=protein&id=73988706)), *Bos taurus* ([NP_001019860.1](https://www.ncbi.nlm.nih.gov/entrez/viewer.fcgi?db=protein&id=67010009)), *Mus musculus* ([NP_001185770.1](https://www.ncbi.nlm.nih.gov/entrez/viewer.fcgi?db=protein&id=311771533)), *Rattus norvegicus* ([NP_476485.1](https://www.ncbi.nlm.nih.gov/entrez/viewer.fcgi?db=protein&id=16924004)), *Gallus gallus* ([NP_001186415.1](https://www.ncbi.nlm.nih.gov/entrez/viewer.fcgi?db=protein&id=313747471)), *Danio rerio* ([NP_001006026.1](https://www.ncbi.nlm.nih.gov/entrez/viewer.fcgi?db=protein&id=54400554)), *Xenopus tropicalis* ([NP_001006836.1](https://www.ncbi.nlm.nih.gov/entrez/viewer.fcgi?db=protein&id=55742005)) revealed that the substituted amino acids in variants p.T16S, p.G78V and p.W140C were highly conserved. The conserved region showing the substituted amino acids alongwith flanking amino acid sequences have been shown in Figure1. (B).

### 3.3 Pathogenic potential of CSRP3 variants

We used *VarCards* (http://159.226.67.237/sun/varcards/) that includes more than 10 *in silico* tools (as listed in materials and methods) to predict the disease-causing potential of identified non-synonymous variants. All these tools predicted the variants p.G78V and p.W140C to be deleterious. However, the missense SNP p.T16S was predicted be deleterious by 6 tools out of 10. The predictions and associated prediction scores of these *in silico* tools have been tabulated in Table1. I-Mutant Suite (http://gpcr2.biocomp.unibo.it/cgi/predictors/I-Mutant3.0/I-Mutant3.0.cgi) was used to predict the stability of protein structure, which revealed that upon mutation, the amino acid changes at 16^th^ (p.T16S) 78^th^ (p.G78V) and 140^th^ (p.W140C) position resulted in decreased stability of CSRP3 protein. This prediction is based on the changes in the free energy between wild type and mutant protein.

### 3.4 Changes in physico-chemical properties of CSRP3 protein due to rare variants

The properties of amino acids, constituting a protein greatly determine its physico-chemical properties, therefore any change in an amino acid at a given position, could potentially lead to changes in the pysico-chemical properties of the protein. These changes can also alter the secondary and tertiary structures of the protein. Hence, to examine the changes in the physico-chemical properties such as hydrophobicity, relative mutability, the percent accessible changes, total beta strand and α-helix of the CSRP3 protein due to variants (p.G78V and p.W140C). We used ProtScale, which predicted significant changes in the hydrophobicity of CSRP3 protein due to both the variants, namely p.G78V and p.W140C as compared to wild type within the region where amino acid substitution has occurred. The percent (%) accessible residues and total beta strand also shown significant changes for both the variants. However, the change in relative mutability was noted in case of p.G78V variant only, no change was observed in variant p.W140C. Similarly, significant changes in α-helix structure was observed as compared to wild type whereas only minor changes were detected in α-helix due to variant p.W140C. The physico-chemical profile of wild type and mutant (p.G78V, p.W140C) CSRP3 protein has been shown in Supplementary Fig. 1

### 3.5 *Prediction of structural changes in* CSRP3 *protein*

PsiPred predicted significant changes in secondary structure of CSRP3 protein due to variants p.T16S, p.G78V and p.W140C (Supplementary Fig. 2 and Figure 2). In case of p.G78V, the structural changes included introduction of a new β-sheet (79-81aa) by replacing an existing coil, introduction of a new helix (141-144 aa) and change in length of β-sheets (16-24aa; 55-58aa) and helix (128-133aa). Besides this, p.W140C variant revealed relatively fewer changes, which include shortening of β-sheet (63-66 aa), and introduction of new β-sheet (127-128aa).

**Figure 2.**
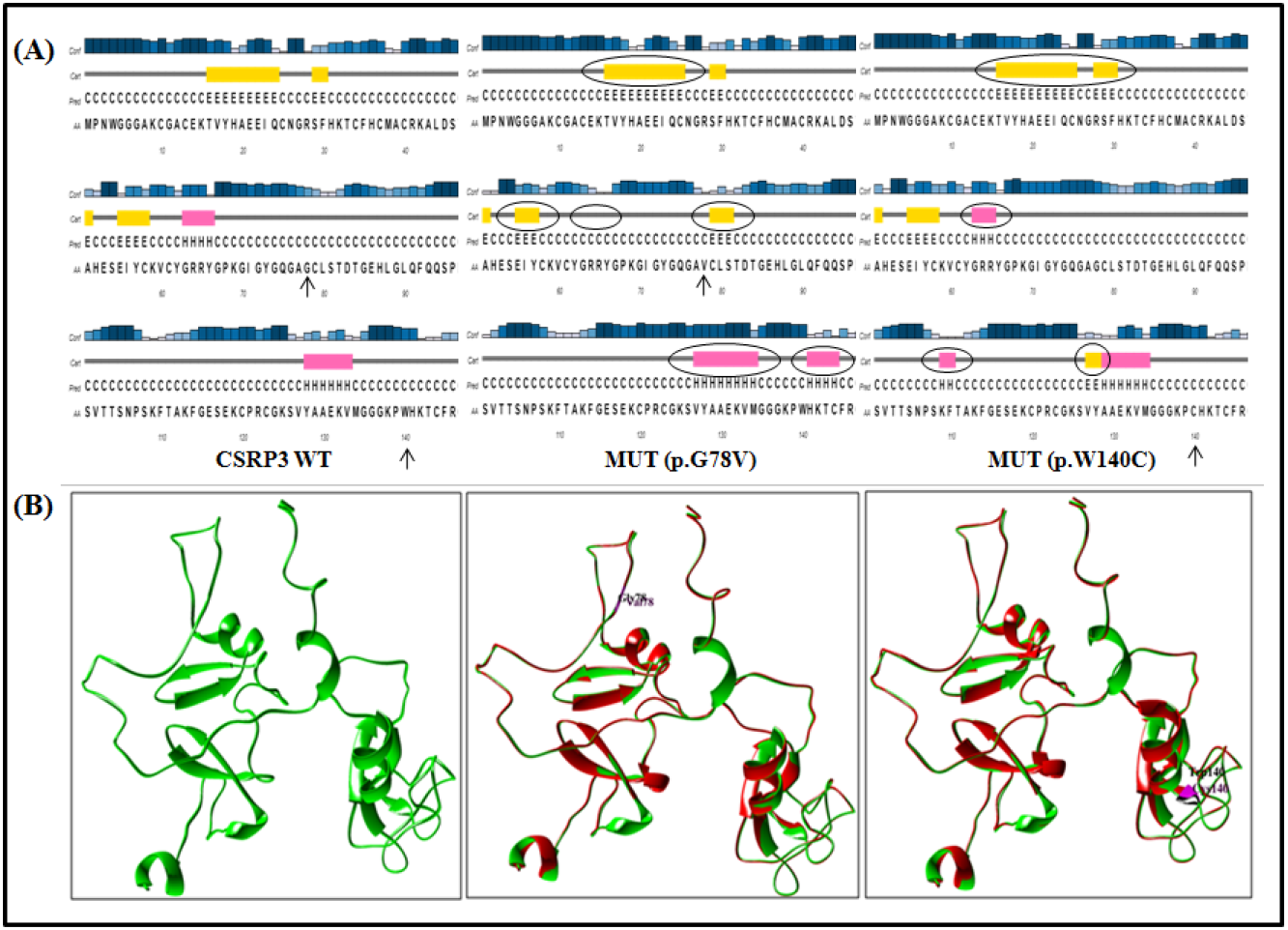
Secondary and tertiary structure based comparison of CSRP3 wild-type (WT) and mutant (MUT) protein by PsiPred and UCSF Chimera respectively. (A) Secondary structure of WT and MUT protein (p.G78V and p.W140C). The position of amino acid changes have been marked with arrow in WT and MUT secondary protein structures and the changes in the MUT structures as compared to wild type have been encircled. (B) Tertiary structure of WT CSRP3 and superimposed three dimensional structures of WT versus MUT (p.G78V; p.W140C) protein using Chimera. WT tertiary structure of CSRP3 protein has been shown in green color while mutant has been shown in red color

The three-dimensional protein structure of both wild type and mutant CSRP3 was modeled which revealed changes in the structure of protein upon mutation as evident by superimposing the wild type and mutant p.G78V or p.W140C protein structure using UCSF Chimera. The visual differences in the wild type and mutant protein structures have been further supported by root mean square deviation (RMSD) values, which is the quantitative representation of similarity or dissimilarity between two or more protein structures. In protein structure prediction, the higher the RMSD values between two structures, the more dissimilar are the two structures and vice-versa. A RMSD value of 0 indicates that two structures those have been superimposed are structurally identical. In our study, the RMSD value for p.G78V was found to be 0.15 Å while for other variant p.W140C the RMSD value was found to be 0.180 Å implicating relatively greater changes in p.W140C mutant than p.G78V mutant CSRP3 protein structure. The aligned structures for each of the mutant versus wild type comparison have been shown in Figure 2.

Interestingly, the SNP p.T16S, also showed significant changes in both secondary and tertiary structure. The superimposed structure of WT CSRP3 and p.T16S mutant protein structure has been shown in Figure 3 A. The WT amino acid threonine (Thr16), which is a part of the zinc finger motif of first LIM domain is involved in making hydrogen bonds with Cys10, Tyr18, and His31 but due to the variant p.T16S, the hydrogen bonding of serine 16 (Ser16) with Tyr10 and His31 is lost (Figure 3. B-C). Also, the RMSD value for these two aligned structures was .11, which is lowest amongst all the three identified missense variants, however, interestingly we observed significant differences in the structure of mutated protein as compared to WT (Figure 3. A).

**Figure 3.**
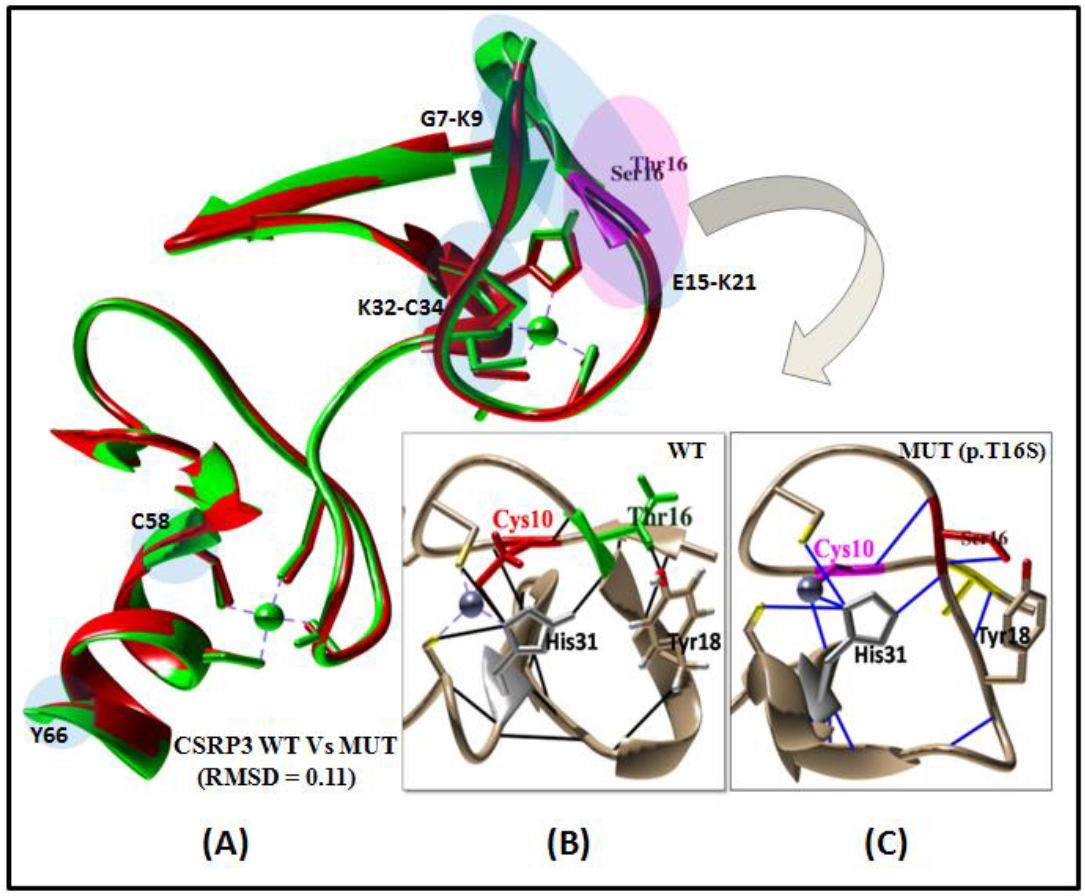
Structural comparison of LIM 1 domain harboring p.T16S variant by UCSF Chimera. (A) Superimposed three dimensional structures of wild type (WT) versus (Vs) mutant (MUT, p.T16S) protein using Chimera. The light magenta colored circle shows the region harboring WT and MUT amino acid residue while the light blue colored circles show the regions of dissimilar conformations across the two structures. The root mean square deviation (RMSD) value of the two superimposed structures is 0.11. WT tertiary structure of CSRP3 protein has been shown in green color while MUT has been shown in red color. (B-C) WT and MUT three dimensional structure showing LIM 1 domain and hydrogen bonding of WT amino acid, threonine (T) and MUT amino acid, serine with neighbouring amino acids.

### 3.9 Impact of non-synonymous variants on the expression and localization of CSRP3 protein

The expression of wild-type and mutantCSRP3 proteins was evaluated by Western blotting, which showed significant differences in the level of protein expression. The level of CSRP3 protein was down-regulated by 1.35 fold (p<0.02) and 1.63 fold (p<0.004) due to the variants p.G78V and p.W140C respectively as compared to wild type (**Figure** 4), which is 26% and 37% reduction in respective expression level compared to wild type.

**Figure 4.**
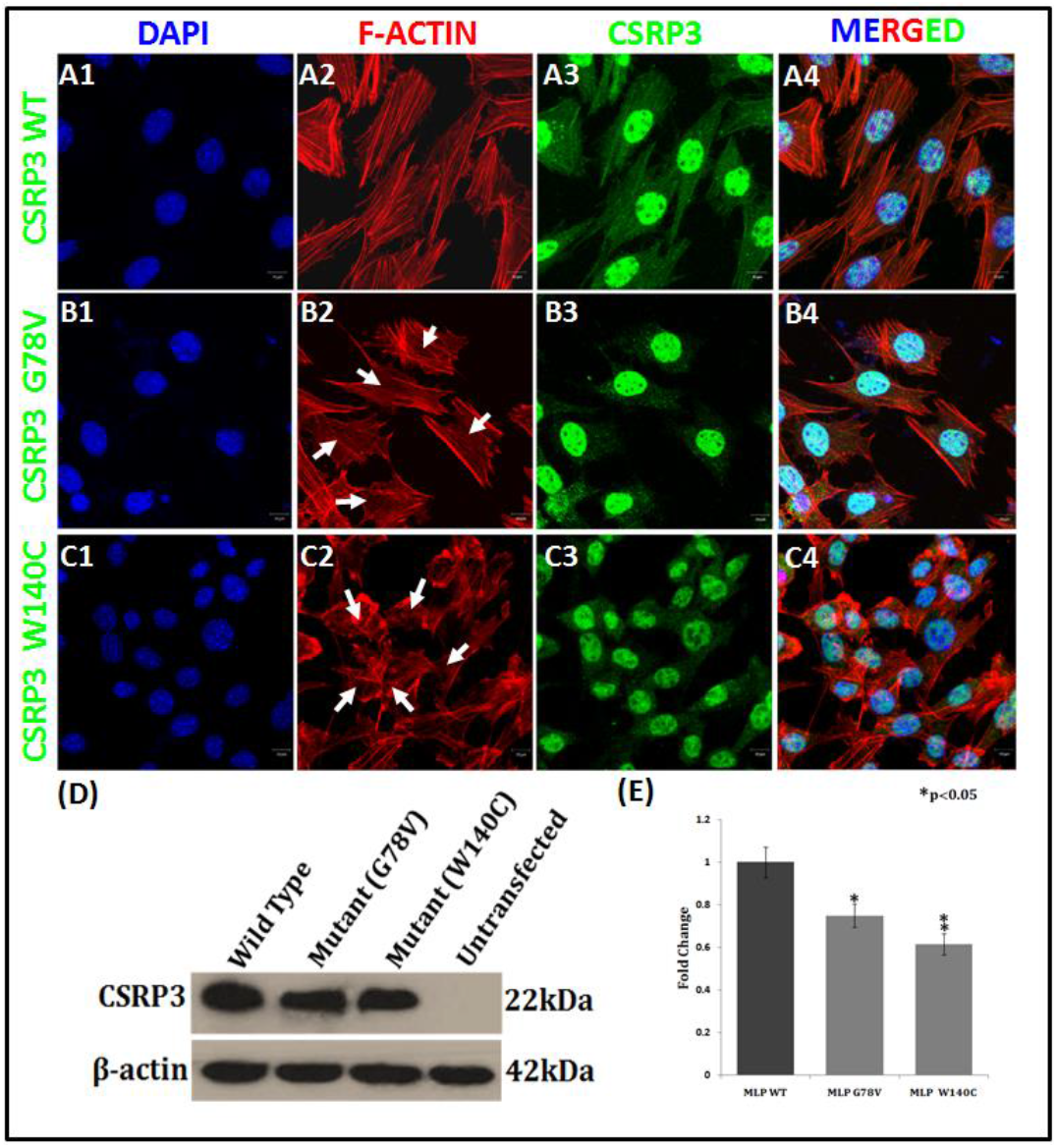
Expression analysis of CSRP3 protein by immunostaining and Western blotting. (A1-C4) Immunostaining in C2C12 cells showing cellular localization of CSRP3-wild-type and mutant protein (p.G78V and p.W140C). (D) Western blot analysis of wild-type and mutant protein showing expression of CSRP3. (E) Representation of protein expression in terms of fold changes, calculated for each of the variant (P>0.05)

The cellular localization of MLP was also studied, by immunostaining, which demonstrated both cytoplasmic and nuclear localization of the protein in wild-type as well as in both the mutants (p.G78V, p.W140C). However, the cytoplasmic expression of CSRP3 protein was more profound in wild-type compared to mutants. There is obvious reduction in cytoplasmic localization of the protein in CSRP3 mutants, which is more pronounced in case of p.W140C mutant. CSRP3/MLP protein is known to act as a scaffold for the regulation of the F-actin based cytoskeleton (Arber et al. 1997; Knoll et al. 2002), therefore we also studied the actin assembly in wild type and mutant protein by Phalloidin staining. Disarrayed actin cytoskeleton was observed in the cells expressing mutant p.G78V and p.W140C proteins when compared to those expressing wild type protein. In case of p.W140C variant, the actin filaments were severely disrupted and the shape and size of cells were also reduced/abnormal (Figure 4. C1-C4).

### 3.10 Effect of CSRP3 variants on the expression of downstream target genes of Z-disc

The endogenous expression of five target genes of *CSRP3* was analyzed by Real time-PCR to evaluate the effect of two non-synonymous variants of CSRP3. The expression of all five targets viz., *Ldb3, Myoz2, Tcap, Tnni3*, and *Ttn*, was significantly reduced in response to both the the variants of CSRP3 (p.G78V and p.W140C) when compared to wild type. The expression of *Ldb3, Myoz2, Tcap, Tnni3, and Ttn*, was reduced by 1.53 (p = 0.00002), 2.08 (p = 0.00001), 1.67 (p = 0.0011), 1.45 (p = 0.0048), and 1.26 fold (p = 0.0012) respectively in response to variant p.G78V. Similarly, the variant p.W140C also caused significant diminution in the expression of above mentioned target genes, viz., 2.04 (p = 0.006), 2.75 (p =0.00000010), 1.8 (p = 0.002), 1.43 (p = 0.008), 1.45 (p = 0.000051) fold reduction in *Ldb3, Myoz2, Tcap, Tnni3, and Ttn* respectively (Figure 5).

**Figure 5.**
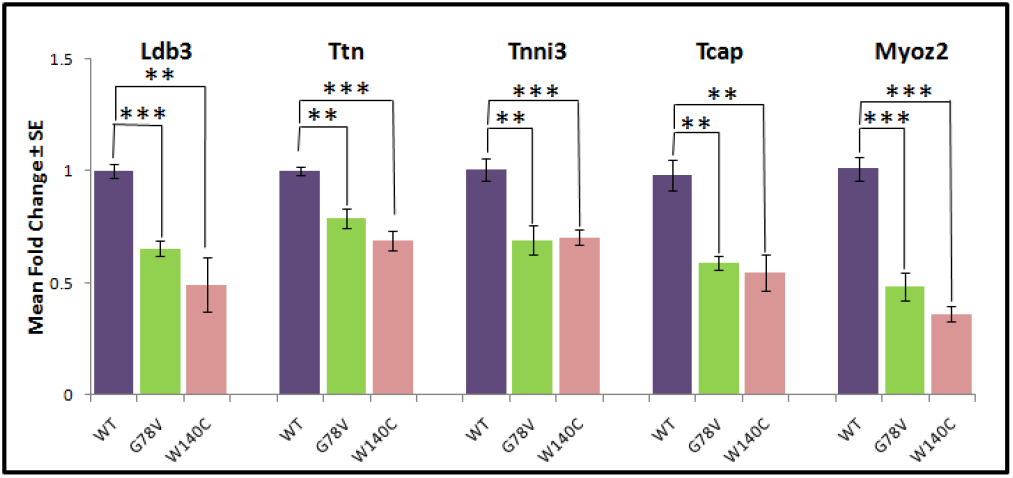
Graph showing results of qRT-PCR assay performed to analyse the expression of other Z-disc genes, i.e. *Ldb3, Ttn, Tnni3, Tcap*, and *Myoz2* due to variants p.G78V and p.W140C, represented as mean fold change ± SE (Standard error).

### 3.11 Impact of non-synonymous variants on CSRP3-ACTN2 protein-protein interaction

In order to evaluate the impact of variants p.G78V and p.W140C on the interaction of CSRP3 protein with ACTN2, GST pull down assay was performed. The binding of ACTN2 with CSRP3 was diminished in case of both the variants(GST-MLP-G78V, GST-MLP-W140C) when compared to (GST-MLP-WT) (Figure 6A).

**Figure 6.**
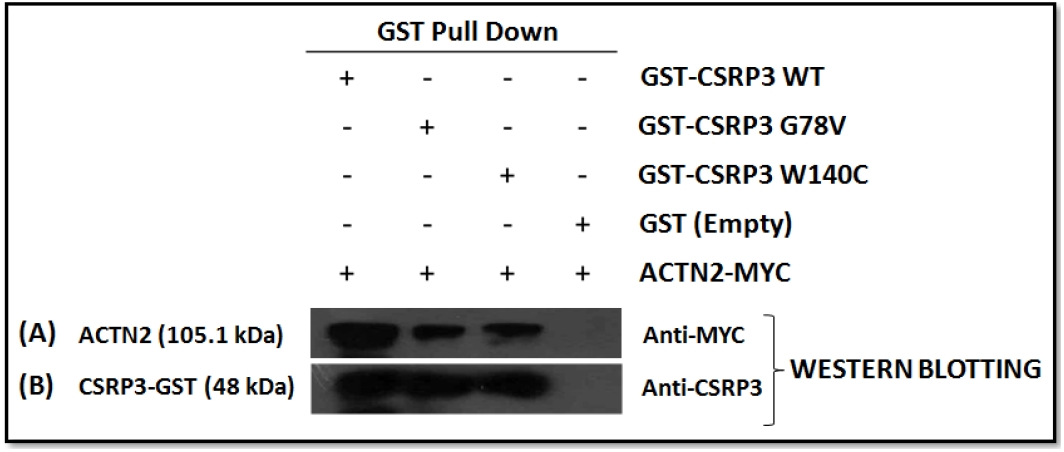
Schematic presentation of GST pull down of ACTN2 by CSRP3 wild-type (WT) and mutants (p.G78V and p.W140C) (A) Detection of ACTN2 pulled down from C2C12 cell lysate transiently expressing ACTN2 using GST-CSRP3 WT (lane 1) and mutant protein (lane 2 and 3 i.e GST-CSRP3 G78V and GST-CSRP3 W140C) immobilized on GST beads with Anti-MYC antibody. (B) Detection of CSRP3 WT and mutant protein in the pulled down fraction using anti-CSRP3 antibody. ACTN2 was not detected in the pulled down fraction with empty GST protein

## 4. Discussion

The CSRP3 is a LIM domain containing protein which functions as a key regulator of striated muscle myogenesis and mutations in the gene can cause both DCM and HCM. The gene encoding the CSRP3 protein is a 20 kb long genomic sequence producing a 0.8 kb transcript organized into six exons encoding a 194 amino acid long protein (Fung et al. 1996 and Knoll et al. 2002) that possesses two Zinc (Zn) finger containing LIM domains, each one having specific binding partners and involved in distinct functional role. In the present study, after screening all six coding exons and exon-intron boundary, we have identified two rare non-synonymous variant and one non-synonymous SNP, those are absent in 100 controls from the same geographical locality. These variants are also absent in other Indian databases such as IndiGen*omes* and INDEX-db that include both North and South Indian population. All three variants are also absent in Genome Asia database. The variants p.T16S and p.W140C are novel while the variant p.G78V (rs963128995) is already reported in dbSNP, however without any MAF and not listed in any other database. The novel non-synonymous polymorphism p.T16S is present in first LIM domain, while the rare variants p.G78V is localized to the first GR module and the p.W140C is confined to the 2^nd^ LIM domain.

The LIM-domain proteins are multifunctional proteins mediating many cellular processes by interacting with other proteins through LIM domain that acts as modular protein binding interface (Schmeichel et al., 1994; Feuerstein et al., 1994). Like other members of CRPs, CSRP3 also shuttles to different sub-compartments of the cell at different stages of development or physiological state. CSRP3 is known to be engaged in Z-disc and act as stretch sensor by interacting with Titin-cap (Tcap) (Knöll et al., 2002), Ankrd1 (CARP) and PKCα (Lange et al.,2016), Calcinurin (Heineke et al., 2005) and Zyxin (Frank and Frey, 2011). Under stress condition, nuclear shuttling of CSRP3 and its interaction with various transcription factors induces gene expression contributing to cellular homeostasis. Although the definite targets for both the LIM domains are not fully understood, depending on the interacting partner, differential role of each LIM domain is predicted. The first LIM domain directly interacts with α-Actinin, a component of Z-disc (Mohapatra et al., 2003; Gehmlich et al., 2004) as well as to the nuclear transcription factor MyoD (Kong et al., 1997) promoting gene expression. On the other hand, the 2^nd^ LIM domain is known to interact with actin cytoskeleton (Arber and Caroni 1996) and to the β-spectrin (Flick and Konieczny 2000). MLP self-dimerizes via N-terminal LIM domain and then interacts with actin cytoskeleton (Hoffmann et al., 2014) through C-terminal LIM domain, thus acting as a scaffold to stabilize the actin filament. It also indirectly interacts with actin filament via alpha-actinin (Louis et al., 1997). In contrast, CSRP3 also regulates depolymerization of F-Actin by interacting with another thin-filament protein Cofilin2 (Papalouka et al., 2009).

In the present study, the first LIM domain variation T16S has been detected in 24 cases (MAF 0.12), although not found in 100 local controls and in none of the existing databases. It localizes within the first zinc finger domain (amino acid 10-58). Though initially bioinformatic analysis using ‘SIFT’ and ‘Polyphen2’ predicted T16S variant as benign, later on while using ‘VarCard’ analysis it become evident that many other tools predict this variant as deleterious (e.g., Mutation Taster, LRT-pred, FATHMM, VEST3, Elgen, GenoCanyon etc.) Considering the later analyses, we further examined the secondary and tertiary structure of this variant and compared the superimposed wild-type versus mutant protein. The secondary structure prediction (Supplementary Figure 2) as well as the superimposed 3D structure (Figure 3.A) have shown significant changes in the confirmation including both the LIM domain, suggesting a debilitating effect of this mutation. However, due to high MAF of this variant, this has not been considered for further *in vitro* functional analysis. Interestingly, this variant is also present in the patient carrying p.W140C mutation with severe phenotype. Despite the fact that this is a polymorphic variation, a possible disease modifier role can’t be ruled out.

Of the two rare missense variants identified in our study, the p.G78V variant possesses a highly conserved glycine residue at 78^th^ position which is replaced by valine. This variant is localized to the first GR domain of the CSRP3 spanning from 63^rd^ through 90^th^ amino acid, which is an important site for protein-protein interactions. Cofilin and α-Actinin are known to bind to CSRP3 through this region (Louis et al 1997; Papalouka et al. 2009). Pathogenic effect of substitution of glycine to valine has been documented in other human diseases for example in Ehlers-Danlos syndrome type IV (Richards et al. 1991; Tromp et al. 1995) & p-glycoprotein (Rao 1995). The variant p.W140C results due to replacement of tryptophan by cysteine at 140^th^ position. It is noteworthy that this is a homozygous variation exhibiting severe disease phenotype. This variant lies in the second LIM domain and most of the bioinformatic tools using VarCard analysis predicted it to be highly deleterious / disease-causing (with a damaging score of 0.91). The second zinc finger domain is also important for protein-protein interactions. Integrin-linked kinase (ILK) and β1-Spectrin interact with CSRP3 via this domain (Flick and Konieczny et al 2000; Postel et al. 2008).

Further, the analysis of physicochemical properties (hydrophobicity, percent accessible residues, relative mutability, total beta strand and alpha helix) of the wild-type when compared to the mutant CSRP3 protein, significant changes have been noted, in case of both p.G78V and p.W140C variants. Glycine being an important residue for proper protein folding, it plays an essential role in the formation of alpha helix. Glycine also has high conformational flexibility since it contains hydrogen as its side chain whereas in case of valine a larger isopropyl group containing side-chain, probably change the protein’s tertiary structure. Similarly, in p.W140C variation the tryptophan is a non-polar aromatic amino acid whereas the substituted cysteine is a sulphur containing residue. These two are the most distantly related amino acids with a Grantham score of 215. The Grantham’s score ranges from 0-215(Grantham 1974). A high Grantham score is considered highly deleterious for amino acid substitutions. Thus, both of these substitutions (p.G78V and p.W140C) possibly lead to changes in stability, folding, structure and conformation of the overall CSRP3 protein. These changes probably can also interfere with the protein-protein interactions of CSRP3 with other interacting partners viz., α-actinin, integrin linked kinase and β1-spectrin (Louis et al. 1997; Flick and Konieczny 2000; Harper et al. 2000; Postel et al. 2008). Furthermore, in case of both p.G78V and p.W140C variants, the secondary structure has changed not only at the site of mutation but also in the other regions of the protein, which suggests that a single amino acid substitution is capable of introducing high degree of conformational changes in the protein. The superimposed 3D protein structures of the wild-type and p.G78V or p.W140C mutants of CSRP3 protein have revealed differences in both the mutants compared to the wild-type, which is further supported by the RMSD values of 0.15 Å and 0.18 Å for p.G78V and p.W140C respectively. These RMSD values indicate that there are remarkable structural changes in both the mutants, albeit p.W140C exhibit relatively higher changes, which is also supported by Grantham score-215, for W to C substitution, implicating that p.W140C substitution is more deleterious as discussed above.

In addition to this, our *in vitro* functional assays evaluating the protein expression in response to variants p.G78V and p.W140C in C2C12 cells, demonstrate reduced levels of CSRP3 protein i.e., 1.35 fold (26%) and 1.63fold (37%) respectively. A decreased level of CSRP3 protein has been reported in case of systolic heart failure (Zolk et al. 2000, Ehsan et al. 2018). Likewise, a missense variant C58G in familial HCM cases has shown a significant reduction (40% reduced) in the levels of CSRP3 protein in myocardium (Geier et al. 2008). MLP also plays an important role in mechano-transduction in heart. Therefore, decreased CSRP3 expression is likely alter the passive mechanical properties of the left ventricle. Omens et al. 2002, have reported that the deficiency of CSRP3 leads to altered passive ventricular mechanics which is associated with ventricular dilation and systolic dysfunction. Our finding of reduced expression of CSRP3 protein due to both the missense variants identified in present study is also supported by the *in sillico* protein stability prediction by I-Mutant, wherein both the variants have been predicted to reduce the stability of the protein. Hence the observed down-regulation of protein is possibly due to less stable mutant protein. Besides, our cellular localization study of wild type and mutant CSRP3 protein has shown both cytoplasmic and nuclear expression in stably transfected C2C12 cells, albeit with remarkable reduction in both mutants, which is more conspicuous in case of p.W140C mutant. The Nuclear Localization Sequence (RKYGPK), present after the first LIM domain, facilitate localization of the protein to the nucleus (Boateng et al., 2007). In differentiating myogenic cells, *CSRP3* has been reported to be initially nuclear, however transported back to cytosol on maturing myotubes and muscle fibers (Arber et al. 1994), thereby shuttling to different sub-compartments such as cytosol, sarcomere, Z-disc, costamere and inercalated disc (Arber and Caroni, 1996; Flick and Konieczny, 2000; Geier et al., 2008; Ehler et al., 2001). In nucleus *CSRP3* acts as a transcription factor interacting with several other transcription factors GATA4, SRF, MyoD and MRF4, thus regulating muscle specific gene expression in cardiac muscles and promoting myogenesis (Buyandelger et al., 2011).

Cytoplasmic function of CSRP3 includes assembly and stabilization of actin cytoskeleton (Hoffmann et al., 2014), which in turn contributes to the maintenance of cytoarchitecture of cardiomyocytes. Depletion of CSRP3 levels exhibit disarray of actin cytoskeleton. Phalloidin staining of C2C12 cells transfected with mutant constructs of both p.G78V and p.W140C have demonstrated highly disorganized cytoskeleton when compared to wild-type. However, the effect of variant, p.W140C is observed to be more severe. Besides disordered cytoskeleton, the size and shape of cells were also irregular. Hoffman and group (2014) have shown that the second LIM domain of CSRP3 is important for its direct interaction with Actin. As described above, the variant p.W140C is located in the second LIM domain of CSRP3 protein, which is recognized as the actin binding interface. Supporting evidences from our *in silico* studies showing distorted 2D and 3D structure of mutated protein with this variation, also suggest that the mutation in this domain led to severe impairment of the actin based cytoskeleton and the cyto-architecture which probably as consequence lead to contractile dysfunction in the heart. To be more precise, when the clinical characteristics of the patient harboring this variant was reviewed, severely impaired left ventricular function with ejection fraction (LVEF) of 16% only, was noted. Due to sudden death of the patient, tissue sample could not be obtained. Moreover, the damaged cytoskeleton observed with the variant p.G78V located in GR domain, could possibly due to improper assembly of the Actin filament indirectly mediated by α-Actinin2, which is a known GR domain binding protein, involved in cross-linking of Actin filament from neighbouring Z-disc (Djinovic-Carugo et al.,1999; Hoffman et al., 2014). In order to test this, we performed GST pull-down assay, which revealed diminished binding of both the mutant (p.G78V and p.W140C) CSRP3 with ACTN2 in case of both the variants thereby predicting destabilization of the actin-based cytoskeleton of the myogenic C2C12 cells. Hence these mutations in CSRP3 protein in DCM cases can lead to diminished protein-protein interactions. The exact mechanism through which this interaction is diminished, requires further investigation, however we speculate that the conformational changes in the CSRP3 protein structure due to the variants p.G78V and p.W140C probably is one of the underlying causes. It has also been reported that ACTN2 binds to 62^nd^-79^th^ amino acids (Harper et al. 2000) of CSRP3, since this binding occurs through a small stretch of amino acids, any change in the amino acid within this aforementioned region may lead to diminished binding of ACTN2 with CSRP3. This fact additionally supports our findings in case of p.G78V. Other studies have also demonstrated decreased protein-protein interactions between CSRP3 and ACTN2 due to genetic variants in *CSRP3* in patients with DCM or HCM (Geier et al. 2003; Mohapatra et al. 2003; Gehmlich et al. 2004).

Aditionally, we studied the effect of variants p.G78V and p.W140C on the expression of certain other Z-dic genes viz., Ldb3, Myoz2, Tcap, Tnni3, and Ttn, those are involved in contractile apparatus. The expression of aforementioned genes are significantly down-regulated due to both the p.G78V and p.W140C variants compared to wild type, albeit variant p.W140C caused significant depletion in expression of Ldb3, Myoz2, Tcap, and Ttn genes compared to G78V. Hence, these genes probably are potential downstream targets, whose expression is possibly regulated by CSRP3. Since all the five genes play an important role in different signaling pathways such as calcium signaling (Yu et al., 2018), myofibrillogenesis, calcineurin signaling (Frey et al., 2004), muscle assembly regulating factor (Markert et al., 2010) therefore altered expression of any of these genes can lead to defective signal transduction in contractile muscle function.

Accumulating evidences from the existing literature (Supplementary Table 1) has claimed that the mutations in *CSRP3* gene are predominantly associated with HCM phenotype (Bos et al., 2006; Geier et al., 2003, 2008; Janin et al., 2018). Nonetheless, several other variants in *CSRP3* namely K69R G72R and W4R are found to be associated with DCM (Knoll et al. 2002, Mohapatra et al. 2003, Hershberger et al. 2018). First ever studied knock-out mouse model for *CSRP3* gene has clearly demonstrated DCM phenotype. However high incidence of HCM cases associated with CSRP3 mutations could be due to limited number of studies performing mutational analysis of CSRP3 gene in DCM cohorts. Lack of large studies with mutigenerational families with DCM would provide a better picture of association of CSRP3 gene with the causation of DCM.

## Conclusion

In a nutshell, our finding of two missense variants in *CSRP3* gene clearly showing association with DCM phenotype, further supported by *in silico* and *in vitro* studies advocate an obligatory role of this gene in patho-physiology of DCM. CSRP3 being a multifunctional protein participate in diverse cellular activity in different sub-cellular compartments such as cytoskeletal organization, mechanical stretch sensing, signal transduction, Ca^+2^ handling and mitochondrial energy metabolism as well as nuclear transcriptional regulation in a context dependent manner. Furthermore, deficiency in CSRP3 results in abnormal Actin cytoskeleton, aberrant cell structure and function possibly giving rise to pathological conditions. Defective binding of CSRP3 to other sarcomeric and Z-disc proteins including α-Actinin, Actin causing destabilization of cyto-architecture as observed in cases of both p.G78V and p.W140C variations. Despite the fact that, the p.T16S is a polymorphic variation, however predicted to have deleterious effects, need further functional characterization to implicate its pathological effect. Although a good number of studies have been carried out, the pathological manifestation (DCM/ HCM) remains intriguing. Extensive investigations, unravelling mechanistic understanding of the overlapping patho-physiology are warranted.

## Data Availability

All data produced in the present study are available upon reasonable request to the authors

## Author’s Contribution

Prerna Giri performed the mutation screening, all the *in silico* and in vitro characterization and prepared the original draft including all the figures and table. Dr. Bhagyalaxmi Mohapatra conceptualized the study, acquired funding, critically supervised and revised the manuscript at every single step. Ritu Dixit performed sited directed mutagenesis. Dr. Ashok Kumar clinically evaluated and diagnosed patients recruited for the study and also helped in collection of blood samples from patients and their available relatives. All authors read and approved the final manuscript.

## Acknowledgements

We are grateful to all the patients, their families and control individuals who participated in the present study. We are extremely thankful to Dr. T. K. Lahiri and Dr. Damyanti Agarwal from Department of Cardio-vascular and Thoracic Surgery, IMS, BHU, Varanasi for their constant support and encouragement in recruitment of patients. We acknowledge University Grants Commission – Centre of Advanced Studies (UGC-CAS) for research fellowship to Ms. Prerna Giri.

