## Supplemental Figure and Table for "Mutations in *CSRP3/MLP*, a Z-disc associated gene are functionally associated with dilated cardiomyopathy in Indian population"

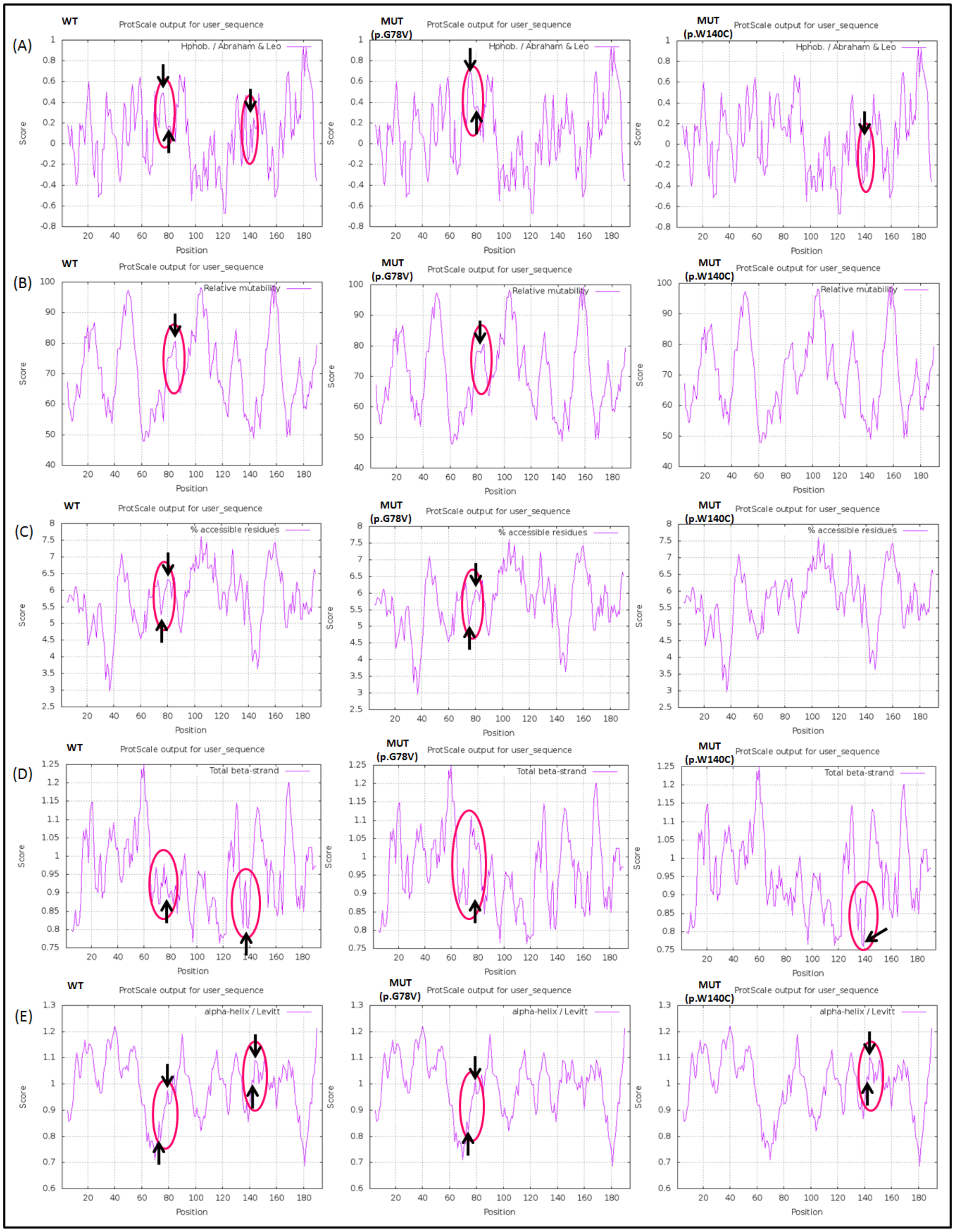

**Supplementary Figure 1. Comparison of physico-chemical properties of wild-type (WT) and mutant (MUT) CSRP3 protein due to variants p.G78V and p.W140C. Representation**

**of hydrophobicity (A), relative mutability (B), the percentage of accessible residues (C), total beta strand (D) and alpha helix residues of WT and MUT CSRP3 protein. The regions showing differences are encircled and marked with arrow. WT = wild-type, MUT = mutant**

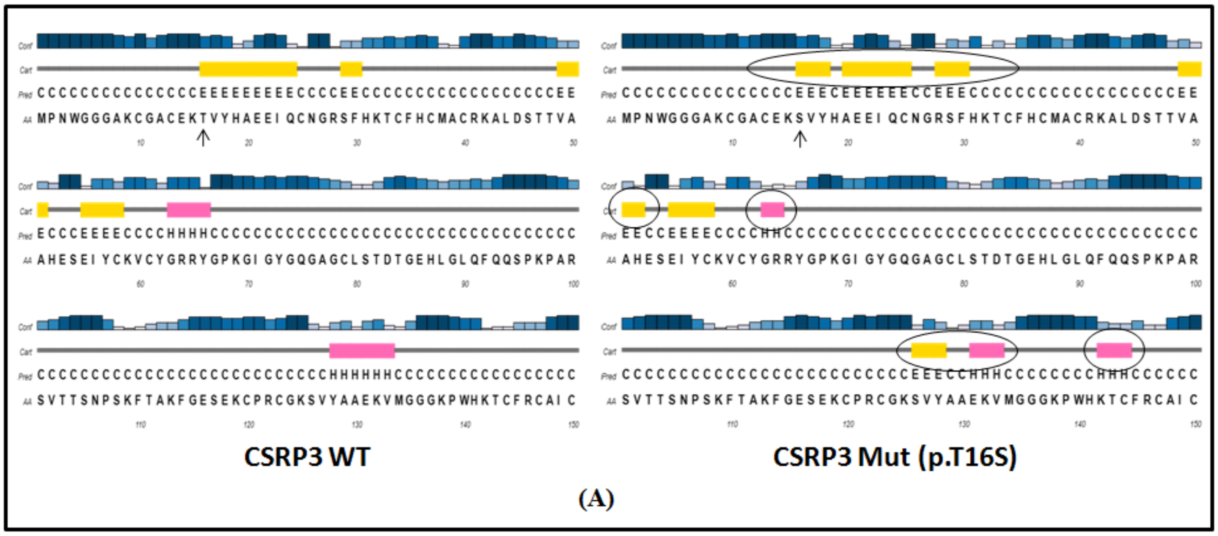

**Supplementary Figure 2. Secondary structure prediction for *CSRP3* p.T16S variant. (A) Secondary structure of WT and MUT protein (p.T16S). The position of amino acid changes have been marked with arrow in WT and MUT secondary protein structures and the changes in the MUT structures as compared to wild type have been encircled.**

**Supplementary Table 1. Table showing the published *CSRP3* variants alongwith the domain of the protein where the mutation is localized, phenotype and the study wherein the variants have been identified.**

| Mutation | Domain | Phenotype | Study |
|---|---|---|---|
| W4R | - | DCM, HCM | Knoll et al. 2002, Mohapatra et al. 2003, Bos et al. 2006, Geier et al 2008, Newman et al. 2005 |
| Y18GInsfsX194 | LIM1 | HCM | Rijsingen et al. 2009 |
| K42/fs165 (K42Gfs) | LIM1 | HCM | Bos et al. 2006 |
| L44A | LIM1 | HCM | Andersen et al. 2008 |
| L44P | LIM1 | HCM | Geier et al. 2003, Bos et al. 2006 |
| S46R | LIM1 | HCM | Geier et al. 2008 |
| S54R/E55G (S54_E55delinsRG) | LIM1 | HCM | Geier et al. 2003 |
| A50T | LIM1 | DCM | Zimmerman et al. 2010 |
| C58G | LIM1 | HCM | Geier et al. 2003, Ehsan et al. 2018 |
| R64C | Gly-rich | HCM | Bos et al. 2006 |
| Y66C | Gly-rich | HCM | Bos et al. 2006 |
| K69R | Gly-rich | DCM with Endocardial Fibroelastosis | Mohapatra et al. 2003 |
| G72R | Gly-rich | DCM | Hershberger et al. 2008 |
| Q91L | - | HCM | Bos et al. 2006 |
| R100H | - | HCM | Andersen et al. 2008 |
| R122Ter | LIM2 | HCM | Lipari et al. 2020 |
| K123Ter | LIM2 | HCM | Janin et al. 2018 |
| K162GInfsX52 | LIM2 | HCM | Janin et al. 2018 |
| T179A | Second Gly-rich | DCM | Zimmerman et al. 2010 |
